# Sampling composition and sequencing effort in the detection of within-hospital Gram-negative genomic clustering: a prospective observational study at two tertiary-care hospitals

**DOI:** 10.64898/2026.09.12.26362918

**Authors:** Leonard Knegendorf, Jelena Erdmann, Karen L. Nielsen, Dmytro Strunin, Rasmus L. Marvig, Dirk Schlüter, Frederik B. Hertz, Susanne Häussler

## Abstract

**Background:** Genomic surveillance of Gram-negative pathogens is generally selective, focusing on suspected outbreaks or resistant isolates. The extent of what such programmes miss remains unknown. It has not been quantified in a large, unselected dataset.

**Methods:** We prospectively whole-genome sequenced isolates of *Escherichia coli, Klebsiella pneumoniae*, and *Pseudomonas aeruginosa* from two tertiary-care hospitals in Copenhagen and Hannover, regardless of resistance phenotype, specimen type, or ward. We analysed 25,888 genomes from 13,712 patients. Clonal clusters were defined by single-linkage clustering at ≤30 SNPs, based on within-patient distances and validated against inpatient movement records. Analyses were patient-based, with hospital comparisons using matched 18-month periods.

**Results:** Clonal clusters included 14.1% of patients in Copenhagen and 16.5% in Hannover. Most clustered patients had no multidrug-resistant isolate (71.3% and 50.0%) and had no intensive care unit (ICU)-associated isolate (86.1% and 68.7%). Sequencing blood-culture isolates alone, the usual trigger for genomic investigation, would have identified only 14 of 900 clustered patients in Hannover (1.6%). Excluding colonisation screening would have missed 240 (26.7%). No targeted strategy detected more clustered patients than random sampling of the same number of isolates. Detection increased with sequencing effort without reaching a plateau. After matching periods and adjusting for ICU association, resistance, and specimen mix, clustering remained more common in Hannover (adjusted odds ratio 1.49, 95% CI 1.28-1.73), suggesting sampling explained much, but not all, of the difference.

**Conclusions:** Most Gram-negative clusters involved non-MDR organisms on general wards and would have been missed by surveillance focused on resistance or intensive care. What is selected determines which patients can be detected; the number of isolates sequenced determines how many are found. The observed 14-16% is a lower bound. Surveillance should sample broadly and representatively, and report what was sequenced and how much.

## Introduction

Gram-negative bacteria are a frequent cause of healthcare-associated infections^1^ and contribute substantially to the global problem of antimicrobial resistance. In 2019, antimicrobial-resistant infections were estimated to be associated with around 4.95 million deaths.^2^ Thus, detecting and preventing the spread of bacteria within and between healthcare facilities is a public health priority.^3^ Whole-genome sequencing (WGS) is increasingly used to investigate possible transmission pathway. ^4–6^ Isolates with genomes that differ at only a small number of single-nucleotide positions are generally considered closely related. Comparing bacterial genomes can help identify previously unrecognised transmission events and support decisions about infection prevention and control.

So far, however, genomic surveillance has mostly been carried out on a selective basis. It is usually triggered by a suspected outbreak or focuses on antimicrobial-resistant isolates. Routine clinical isolates and susceptible colonising organisms are rarely included. Even large multicentre studies have generally been limited to antimicrobial resistance and single-species analysis,^7^ whereas some single-centre studies have examined broader isolate collections from intensive care units (ICUs) or routine clinical specimens.^8,9^ Studies including both resistant and susceptible colonising isolates have likewise often remained restricted to specialised wards.^10^

What surveillance reveals depends on where specimens come from, which types are collected, and how many are sequenced. A hospital that sequences colonisation swabs may detect transmission clusters that would remain hidden in a hospital sequencing only blood cultures. Comparisons between the two settings—and benchmarks based on them—may therefore mistake differences in sampling for differences in transmission. Few studies have quantified this effect.^11,12^ Doing so requires a sequencing strategy that does not selectively target particular specimens. It is also still unclear what proportion of a hospital’s isolates needs to be sequenced before the number of detected clusters reaches a plateau.

How much sampling affects the detection of transmission is likely to differ between species. Different species occupy different ecological niches and vary in their ability to colonise and persist in patients. This may be particularly relevant for Klebsiella *pneumoniae*, which can establish clonal gut carriage and spread within and between healthcare networks.^7,13^ As a result, the types of specimens included in genomic surveillance may have a greater impact for some species than for others.

In this study, we conducted prospective, large-scale WGS of *Escherichia coli, K. pneumoniae* and *Pseudomonas aeruginosa* at two tertiary-care university hospitals in Denmark and Germany. These three species are among the most clinically important Gram-negative pathogens in healthcare-associated infections. Clinical isolates were included irrespective of resistance phenotype or specimen type; screening isolates followed site-specific policies. The resulting dataset comprised 25,888 genomes and provides an unusually broad basis for describing hospital clustering as it appears when nothing is filtered out in advance.

Using a uniform single-nucleotide polymorphism (SNP)-based clustering, which we validated against patient-movement data, we asked which patients and which organisms are involved in detectable within-hospital clustering, and how much of that clustering different surveillance strategies would find. A companion study examines clones that are shared across countries in the same cohort. Here, we focus on clustering within hospitals and on how sampling shapes what is detected.

## Methods

### Study design and isolate collection

In this prospective observational study, routine diagnostic isolates of *E coli, K. pneumoniae* sensu stricto, and *P. aeruginosa* were sequenced at Rigshospitalet, Copenhagen, Denmark, and Hannover Medical School, Germany, irrespective of resistance, specimen type, or ward. Collection covered November, 2019, to December, 2023, in Copenhagen and March, 2021, to August, 2022, in Hannover. All isolates of these species identified by the two clinical microbiology laboratories during these periods were eligible, and no further selection was applied. Where a sample yielded more than one isolate of the same species, Hannover retained every isolate undergoing susceptibility testing, whereas Copenhagen generally retained one per patient, day, and specimen type, with additional isolates from invasive infections. Screening isolates were included irrespective of resistance in Hannover but restricted to carbapenem-resistant organisms in Copenhagen. Reporting follows Strengthening the Reporting of Molecular Epidemiology for Infectious Diseases (STROME-ID).^14^ Most Copenhagen isolates were also included in an earlier study that analysed these and further species to model the economic and health impact of integrated genomic surveillance.^15^

### Ethics and consent

Isolate collection and analysis complied with relevant laws and institutional guidelines. The study was approved by the ethics committee of Hannover Medical School (No. 10372_BO_K_2022), the Regional Danish Patient Safety Authority (R-21015888), and the local data-protection agency (Pactius P-2020-743). The need for informed consent was waived by the responsible authorities at both sites.

### Genomic and statistical analysis

Sequencing, quality control, and SNP-distance calculation are described in the Supplementary Methods. Candidate isolate pairs were pre-grouped by gene content (Jaccard similarity ≥0.80) before pairwise SNP distances were calculated. Primary single-linkage components used eligible relationships at ≤30 SNPs across both hospitals. A local cluster was defined as a hospital-specific cluster of a given species comprising at least two patients. In overall patient-level analyses, each patient was counted once within each hospital. In species-specific contact analyses, the unit of analysis was the patient-species-cluster combination, such that a patient could contribute more than once if assigned to multiple species or clusters. Cross-hospital clusters were not considered. Threshold and chaining sensitivity analyses are reported in Tables S4A-B and S7.

Screening isolates were classified as screening isolates regardless of the specimen type; remaining categories represented clinical, non-screening isolates (Table S2), Patients could contribute to several categories. Multidrug-resistant (MDR) classification used an adaptation of the Magiorakos framework based on routine susceptibility results,^16^ with one laboratory-reported exception. Susceptibility testing used disk diffusion in Copenhagen and minimum inhibitory concentration determination in Hannover, on reduced and partly site-specific panels, so MDR status is not strictly comparable between hospitals (Supplementary Methods), ICU association indicated any isolate sampled on an ICU ward.

Patient-level logistic models examined local cluster membership using hospital, ICU association, MDR status, and specimen indicators. Analyses covered the full cohorts and two separate sensitivity analyses: restriction to the Hannover calendar window and patient-number matching using 1000 random subsamples of the full Copenhagen cohort (Tables S6A-B), Random subsampling varied isolate or patient numbers. For depth analyses, we retained one random isolate per patient and added predefined fractions of the remaining isolates. Separately, the sampling strategies used in Hannover were compared with random subsets of the same size. Network components and relevant patient covariates were reconstructed after selection. Simulation settings, diversity methods, and handling of the other hospital’s isolates are described in the Supplementary Methods.

### Epidemiological analysis

Inpatient-movement records identified direct or indirect ward contact with a cluster co-member, using identical definitions at both hospitals. Direct contact required overlapping recorded stays on the same ward; for indirect contact, the stay boundaries were extended by 14 days. Stays beginning on or after the later of the two patients’ first cluster-isolate sampling dates were excluded. Overall estimates counted each patient once. Unavailable movement data were treated as no documented link; complete-case analyses retained patients with data for at least one assignment (Table S5A), We also assessed clusters with any documented contact (Table S5B), Proportions have Wilson 95% confidence intervals (Cis), A separate proxy, defined as isolates from two patients sampled on the same ward less than 14 days apart, was evaluated by 1,000 within-hospital, within-species permutations of cluster labels, with one-sided empirical *p* values and a plus-one correction (Table S5D).

## Results

### An unselected, highly diverse Gram-negative population

To capture the routine hospital population of three clinically important Gram-negative pathogens— *E. coli, K. pneumoniae* and *P. aeruginosa—*we sequenced routine diagnostic and colonisation-screening isolates collected at two tertiary-care hospitals: Rigshospitalet in Copenhagen and Hannover Medical School. At both hospitals, clinical isolates were included without selection by resistance phenotype, ward of origin, or specimen type. Hannover applied the same unselected approach to screening isolates, whereas Copenhagen sequenced screening isolates only when carbapenem-resistant (Methods), In total, we sequenced 25,888 genomes: 15,383 *E. coli*, 4,734 *K. pneumoniae* and 5,771 *P. aeruginosa* isolates from 13,712 patients. The Copenhagen dataset comprised 13,248 isolates from 8,262 patients collected over 50 months, while the Hannover dataset included 12,640 isolates from 5,450 patients collected over 18 months (Table SI),

All three populations were highly diverse, with no single clone predominating at either hospital (Figure SI), Each species included several hundred sequence types (STs) at both sites, and Gini-Simpson diversity was consistently high, ranging from 0.96 to 0.98. No individual ST accounted for more than approximately 11% of any species. *E. coli* ST131 was the most common lineage at both hospitals, representing about 10% of isolates, consistent with its globally disseminated status.^17^ In Hannover, the most frequent *K. pneumoniae* lineage was ST86, which formed one large hospital cluster (see below).

Copenhagen had more distinct STs than Hannover, as expected from its larger sample size and longer sampling period. The difference was no longer apparent once patient numbers were matched, which in fact more than compensated for it. Gini-Simpson diversity and richness estimates were similar across all comparisons (Figure S1E-G; Table S3),

### A uniform, data-driven clustering threshold

We grouped closely related isolates into genomic clusters and derived the clustering threshold from the data. For each species, we compared the minimum pairwise SNP distance observed among isolates from the same patient with that observed between isolates from different patients. The analysis was restricted to isolate pairs collected within 60 days and differing by no more than 120 SNPs (Figure S2).

Most within-patient comparisons were within 30 SNPs: 95.6% for E, *coli*, 96.7% for *K. pneumoniae* and 86.2% for *P. aeruginosa*. Distances between isolates from different patients were generally much larger. However, many of the pairs separated by 30 SNPs or fewer also came from different patients—for example, 43% for *K. pneumoniae* (Figure S2E), These closely related isolates from different patients are compatible with transmission, but do not establish it.

We therefore defined clonal clusters using a single-linkage threshold of 30 SNPs or fewer. We deliberately chose a permissive cut-off to avoid excluding isolates that could plausibly belong to the same cluster. It provided the best fit for *E. coli* and *K. pneumoniae*, for which the 95th percentiles of within-patient distances were 27 and 25 SNPs, respectively. The fit was less satisfactory forP. *aeruginosa*, whose 95th percentile reached 64 SNPs; this species is therefore likely to be underclustered at the 30-SNP threshold. We retained the same cut-off for all three species to allow direct comparisons.

We assessed the robustness of the threshold in two ways. First, the difference between the hospitals was not a consequence of the selected cut-off. A greater proportion of patients clustered in Hannover than in Copenhagen even when we applied stricter thresholds of 20 SNPs or fewer (13.5% vs 11.2%) and 15 SNPs or fewer (11.7% vs 9.1%, Table S4A), Second, we examined whether single-linkage clustering had produced artificial chains. Of the 728 local clusters containing isolates from at least two patients, 574/728 (78.8%) had all pairwise distances within the 30-SNP threshold and 33/728 (4.5%) contained a pair above 120 SNPs. Table S4B shows single-linkage chaining analysis stratified by species. Increasing the threshold to 70 SNPs for P. *aeruginosa* increased the number of clustered patients from 238 to 419 in Copenhagen and from 204 to 392 in Hannover (Table S7),

### Epidemiological links support the clustering threshold

For each clustered patient, we assessed whether another member of the same cluster had stayed on the same ward during an overlapping admission (direct contact), within a 14-day window around it (indirect contact), or not at all (Figure 1; Methods) .

**Figure 1.**
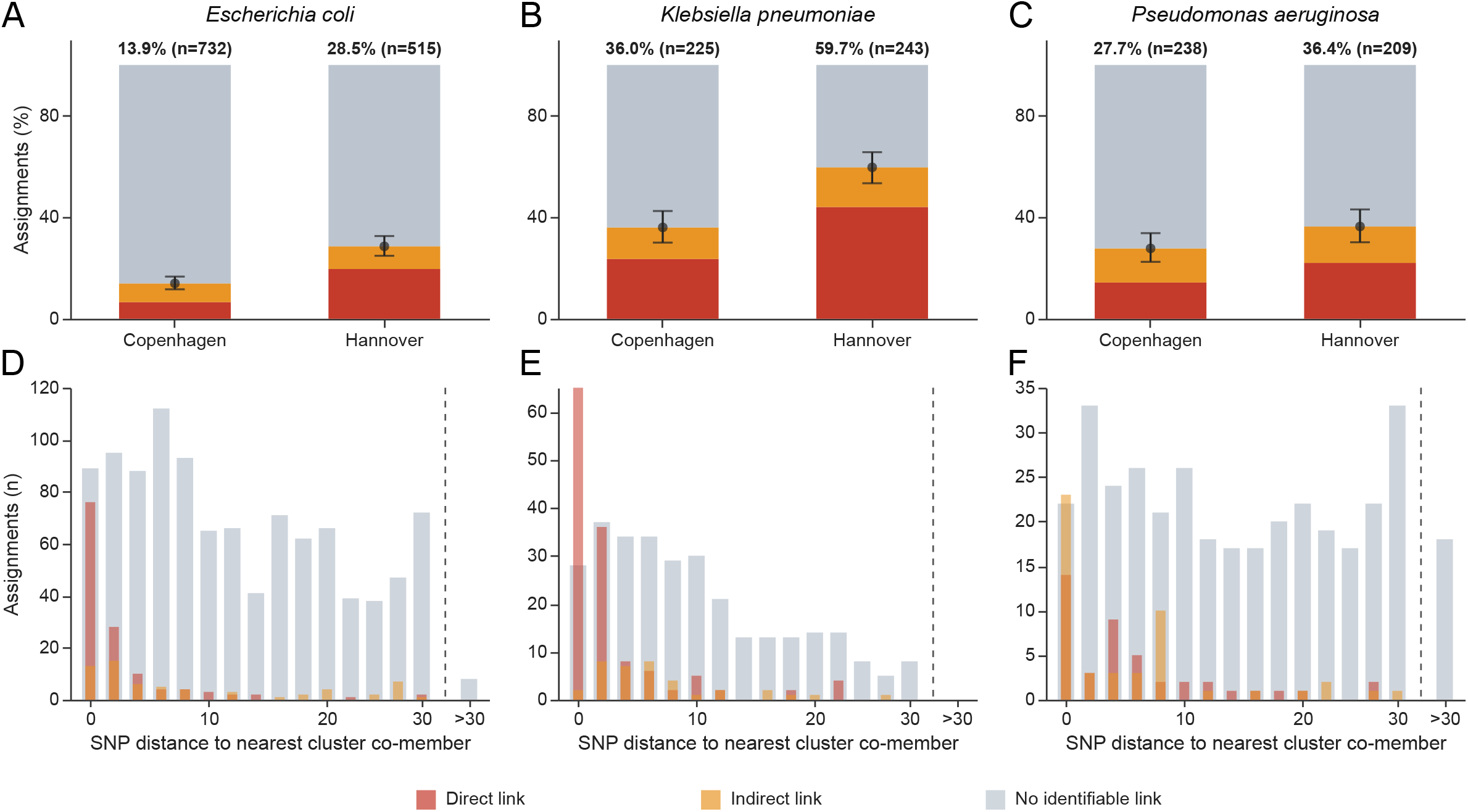
Epidemiological links among patient-species-cluster assignments. (A-C) Proportions of assignments with direct, indirect, or no documented ward contact *in Escherichia coli, Klebsiella pneumoniae*, and *Pseudomonas aeruginosa*, respectively, by hospital. Labels and error bars summarise any documented direct or indirect contact with a cluster co-member and its Wilson 95% confidence interval; *n* denotes the total number of assignments in each hospital-species group. (D-F) Minimum SNP distance to an isolate from another patient in the same local cluster for the corresponding species, pooled across hospitals and classified by the strongest documented link to a tied nearest genomic co-member. Distances up to 30 SNPs are displayed in 2-SNP bins; larger nearest distances are grouped separately. A patient may contribute to more than one species or cluster. No identifiable link includes unavailable movement data. SNP=single-nucleotide polymorphism.

Documented director indirect ward contact with a cluster co-member was identified for 243 of 1,161 clustered patients in Copenhagen (20.9%, 95% CI 18.7-23.4) and 335 of 900 in Hannover (37.2%, 34.1-40.4; Table S5A), Patients without available inpatient-movement data (57 of 1,161 in Copenhagen and 125 of 900 in Hannover) were retained as having no documented link. Excluding them increased these proportions to 22.0% and 43.2%, respectively, an increase of 1.1 and 6.0 percentage points. The higher documented-contact proportion in Hannover therefore persisted.

At least one contact was documented in 92 of 388 local clusters in Copenhagen (23.7%, 95% CI 19.8–28.2) and 131 of 340 in Hannover (38.5%, 33.5-43.8; Table S5B), Links were concentrated at short genomic distances for *E. coli* and *K. pneumoniae*, whereas direct contacts in *P. aeruginosa* occurred across a broader distance range (Figure 1D-F; Table S5C), A separate sampling-event ward-contact analysis showed higher observed contact proportions than after random reassignment of cluster labels in every hospital-species stratum (all p=0.001, the lowest value attainable with 1,000 permutations; Table S5D),

### Most clustering involves non-MDR organisms outside intensive care

Overall, 14.1% of patients in Copenhagen and 16.5% in Hannover belonged to a local cluster. Although MDR isolates were more common among clustered than non-clustered patients, 71.3% of clustered patients in Copenhagen and 50.0% in Hannover had no MDR isolate. Overall, most patients were not treated in intensive care, and consequently most clustered patients came from outside the ICU. The proportion of clustered patients with no ICU-associated isolate was 86.1% in Copenhagen and 68.7% in Hannover (Figure 2; Table SI), Nearly all clustered patients in Copenhagen and most in Hannover also had at least one routine clinical, non-screening specimen sequenced.

**Figure 2.**
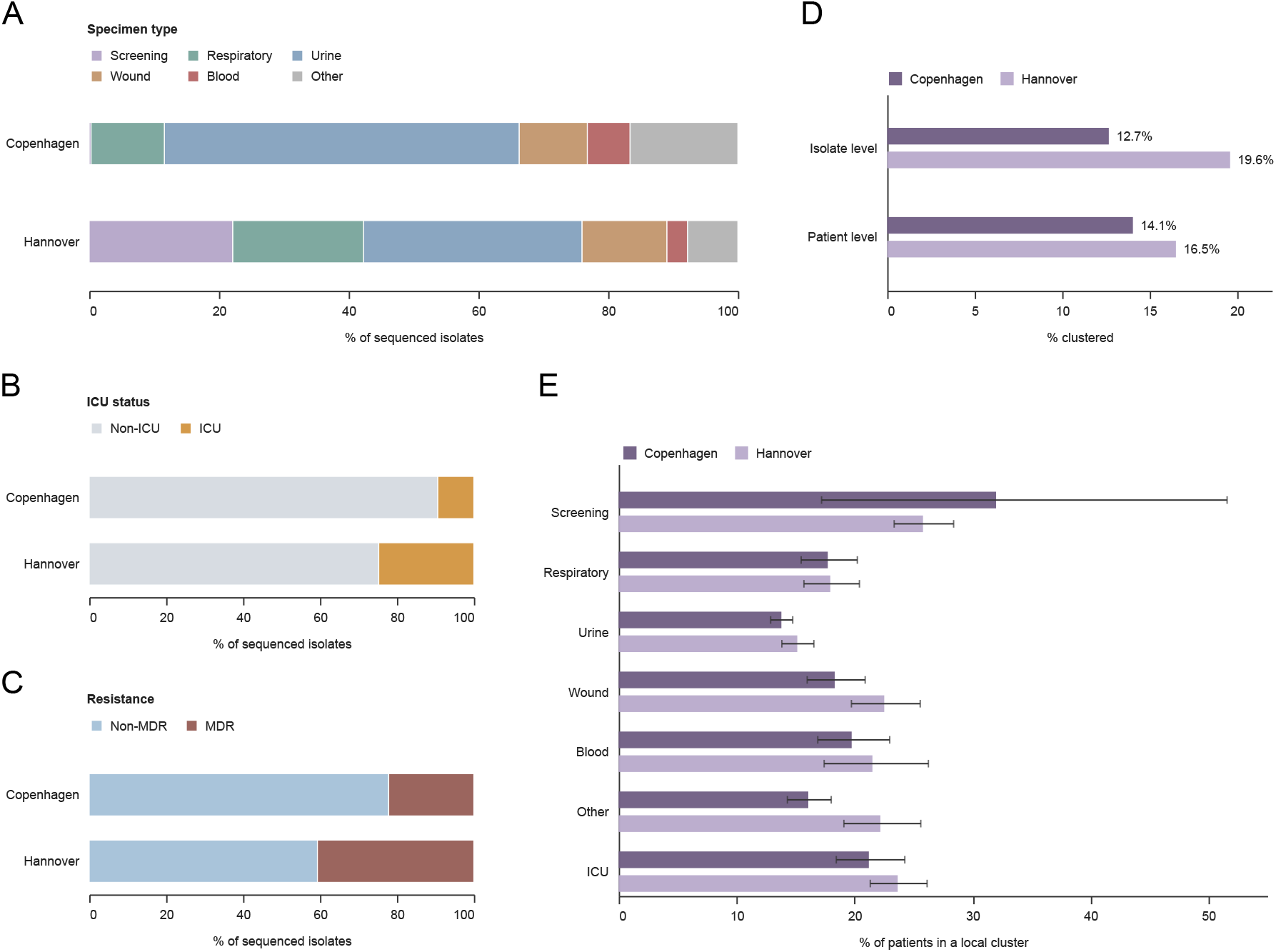
What is sequenced shapes apparent clustering. (A-C) Composition of sequenced isolates by specimen type, ICU association, and MDR status, respectively, at each hospital. Screening status overrides specimen type, so all remaining specimen categories represent clinical, non-screening isolates. (D) Proportions of isolates and unique patients belonging to local clusters. (E) Proportions of patients belonging to local clusters among patients with at least one isolate in the respective specimen category or with an ICU-associated isolate, with Wilson 95% confidence intervals. Patients may contribute to multiple categories through different isolates but are counted once per category and hospital. Catheter and drain specimens are included in other. ICU=intensive care unit. MDR=multidrug-resistant.

### What is sequenced shapes what is detected

Both hospitals sequenced clinical isolates from all three species, but the specimens included in the two datasets differed substantially (Figure 2A-C), Urine and blood specimens made up a larger share of the Copenhagen dataset and respiratory specimens a larger share of the Hannover dataset. The largest difference concerned colonisation screening, which accounted for 22.1% of isolates in Hannover but only 0.3% in Copenhagen (Figure 2A), Hannover also contributed larger proportions of ICU-associated and MDR isolates (24.7% vs 9.4% and 40.6% vs 22.1%, respectively; Figure 2B-C; Table SI),

### Cluster detection increases with sequencing effort

Both the number of local clusters and the proportion of patients assigned to a cluster increased as more isolates were sequenced (Figure 3A-B), When sampling was based on 25% of the patients, 6.4% and 7.9% of patients, respectively, belonged to a cluster. With the full datasets, these proportions increased to 14.1% in Copenhagen and 16.5% in Hannover.

**Figure 3.**
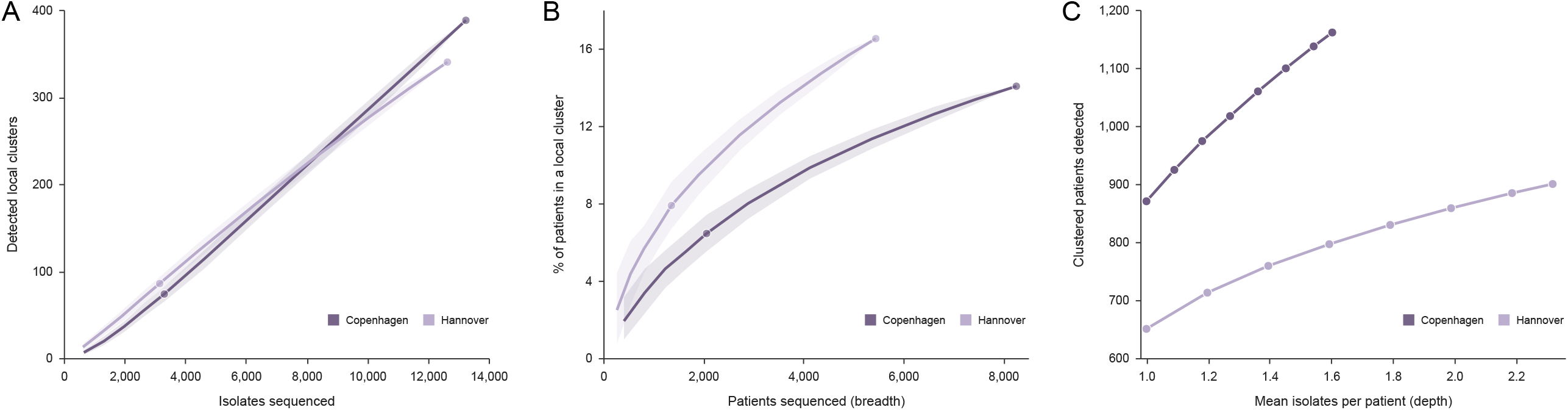
Detected genomic clustering increases with sequencing effort. (A) Number of reconstructed local clusters as increasing numbers of isolates are sampled. (B) Proportion of sampled patients belonging to a local cluster as patient coverage increases, retaining all isolates of selected patients. (C) Number of clustered patients detected as mean isolates per patient increases, starting with one randomly selected isolate from every patient and adding random fractions of the remaining isolates. Estimates are means across 500 draws; shaded bands in A and B show the 10th-90th percentiles. Full-data endpoints are evaluated directly. Components are reconstructed after each subsample, with the other hospital’s isolates retained as a fixed genomic background. Local detection requires at least two patients at the hospital being evaluated; separate qualifying fragments are counted separately in A. Neither rarefaction curve reaches a plateau within the observed range.

In the full cohorts, the crude odds ratio (OR) for patient-level cluster membership in Hannover versus Copenhagen was 1.21 (95% CI 1.10-1.33), falling to 0.96 (0.86-1.07) after adjustment for ICU-associated isolates, MDR status, and specimen categories. Screening had the strongest adjusted association among the included indicators (adjusted OR 2.22,1.84-2.67), However, restricting both datasets to the Hannover calendar window left 3,229 patients in Copenhagen and 5,450 in Hannover. The adjusted hospital OR was then 1.49 (95% CI 1.28-1.73), Separately, matching patient numbers across 1,000 random samples gave a median adjusted OR of 1.23 (median model-based 95% CI 1.08–1.39; Table S6A), Hospital comparisons therefore depended on the observation period, patient coverage, and measured sampling composition. Model-specification sensitivity analyses are reported in Table S6B.

Additional isolates from already included patients also increased detection, consistent with patients carrying more than one lineage (Figure 3C), Neither the isolate-based nor patient-based rarefaction curve reached a plateau within the observed range. The full-data proportions therefore represent the clustering detectable under the observed sampling, rather than a complete measure of hospital transmission.

### *K. pneumoniae:* screening for carriage changes what is detected

In Hannover, 22.2% of patients with *K. pneumoniae* belonged to a local cluster, compared with 14.4% in Copenhagen. Of 237 clustered patients in Hannover, 99 (41.8%) had a screening isolate and 15 (6.3%) a blood-culture isolate; only three of 223 clustered patients in Copenhagen (1.3%) had a screening isolate (Figure 4A). One Hannover ST86 cluster included 13 patients over 11 relative months. Of its 174 isolates, 169 were screening isolates and five were clinical isolates; blood cultures identified the clone in three patients (Figure 4B). This illustrates how screening can reveal carriage-associated clustering that is poorly represented by blood cultures. It does not establish a higher transmission rate in Hannover.

**Figure 4.**
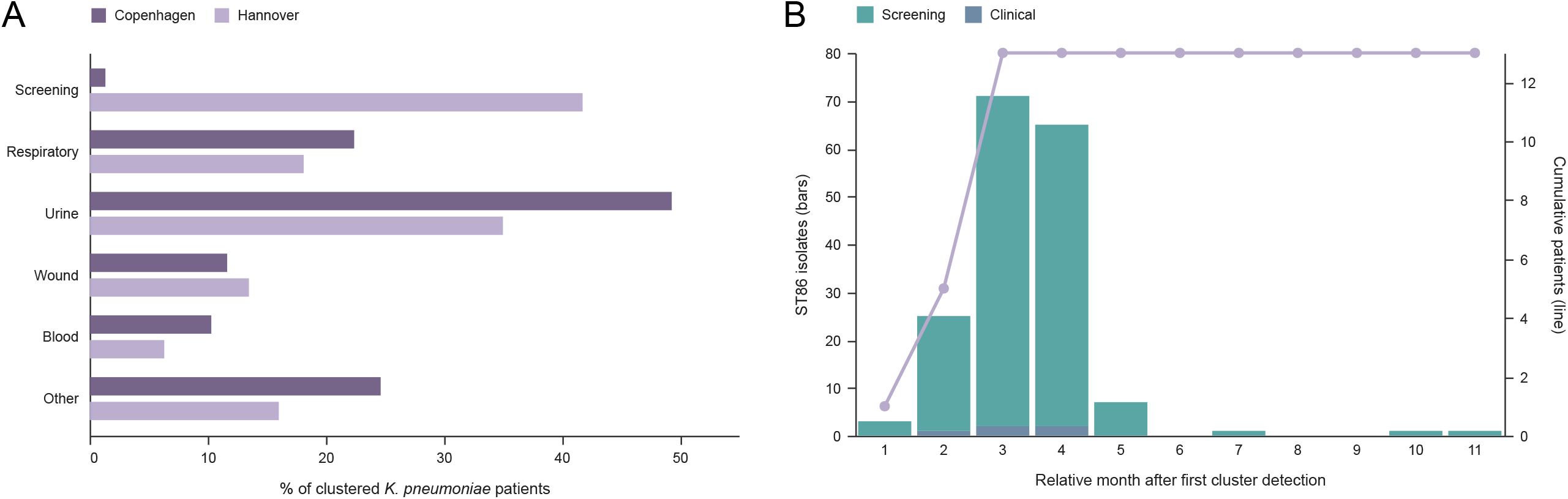
Contribution of screening to *Klebsiella pneumoniae* cluster detection. (A) Proportion of clustered *K. pneumoniae* patients with at least one isolate in each analytical specimen category, by hospital. Screening overrides specimen type; patients may contribute different isolates to several categories. (B) Monthly detection of the Hannover ST86 cluster, comprising 174 isolates from 13 patients. Bars show screening and clinical isolate counts, and the line shows the cumulative number of detected patients. Relative months are measured from the first cluster detection. ST=sequence type.

### No targeted strategy outperformed unselected sequencing of the same size

We recalculated genomic clusters within targeted isolate subsets from Hannover and compared detection with random samples containing the same number of isolates (Figure 5A). Sequencing 405 blood-culture isolates identified 14 of 900 clustered patients, compared with a random-sampling median of 17 (2.5th-97.5th percentile 9-27). Similarly, sequencing 2,796 screening isolates identified 185 clustered patients, compared with 195 (167-219) after size-matched random sampling. Both strategies fell within the corresponding central 95% random-sampling ranges; the remaining targeted strategies fell below them.

**Figure 5.**
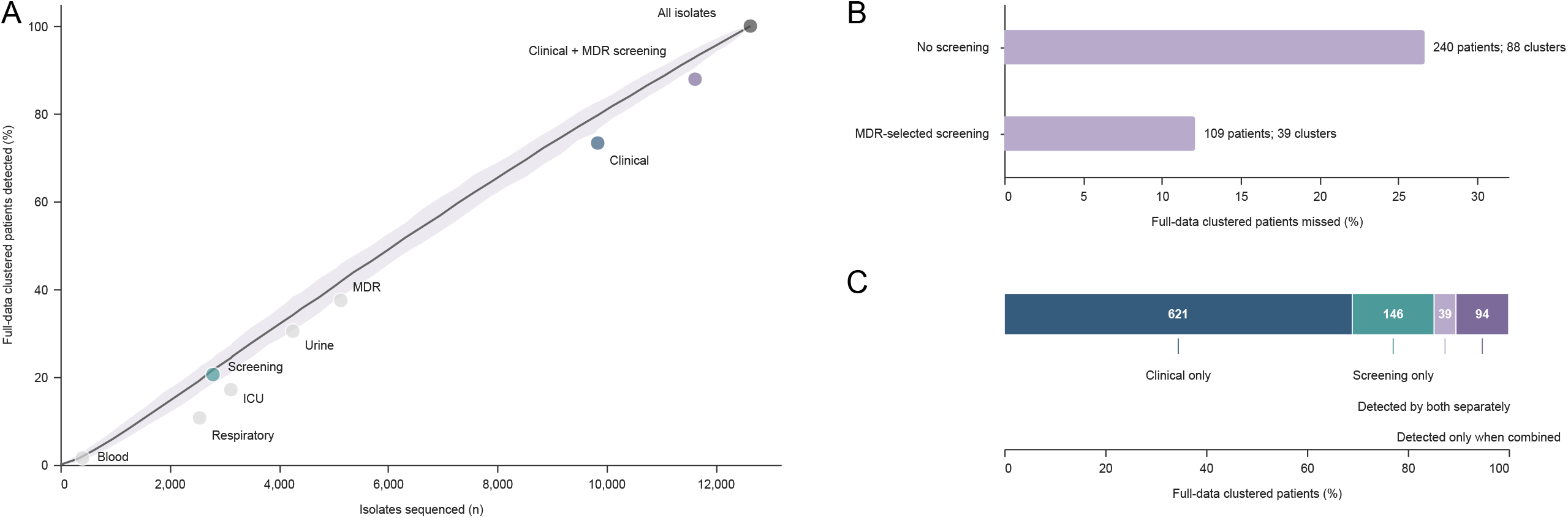
Consequences of targeted sequencing strategies in Hannover. (A) Percentage of the 900 full-data clustered patients detected at each strategy’s isolate budget. The grey line and shaded band show the median and 2.5th-97.5th percentiles of 500 random samples at the corresponding isolate budgets. Components are reconstructed for each subset, retaining Copenhagen isolates as a fixed genomic background. Specimen-type strategies include clinical, non-screening isolates only. (B) Percentage of full-data clustered patients missed after omitting all screening isolates or retaining only MDR screening isolates while keeping all clinical isolates. Labels also report the numbers of undetected reference local clusters. MDR-selected screening is not an exact reconstruction of Copenhagen’s carbapenem-resistance-based policy. (C) Mutually exclusive patient groups detected by clinical-only analysis, screening-only analysis, both analyses independently, or only when both isolate sets are analysed together. MDR=multidrug-resistant.

Retaining all clinical isolates but sequencing only MDR screening isolates would omit 1,013 isolates (8.0%), leaving 39 local clusters undetected and missing 109 clustered patients (12.1%; Figure 5B). Of these missed patients, 82.6% had no MDR isolate. Excluding all 2,796 screening isolates (22.1%) would leave 88 local clusters undetected and miss 240 clustered patients (26.7%). In both scenarios, the proportion of clustered patients missed exceeded the proportion of isolates omitted.

Of the 900 clustered patients, 621 were detected by clinical-isolate analysis only, 146 by screening-isolate analysis only, and 39 by both analyses independently. A further 94 were detected only when both isolate sets were analysed together (Figure 5C).

## Discussion

We prospectively sequenced isolates from three major Gram-negative pathogens at two tertiary-care hospitals, without selecting clinical isolates by resistance or specimen type; screening-isolate inclusion differed between hospitals. A single, data-derived threshold of 30 SNPs or fewer produced clusters that were supported by patient-movement data. Most of the clustering we detected involved non-MDR isolates in patients with no ICU-associated isolate. Surveillance directed at resistant organisms or at intensive care would therefore miss the larger part of the signal.

The extent to which this signal is captured depends on which isolates are sequenced. Cluster membership was associated with specimen type, intensive-care exposure, and antimicrobial resistance, but the strongest association was with colonisation screening. Sequencing triggered by blood cultures—the conventional starting point for genomic investigations—would have detected only 1.6% of clustered patients in Hannover, and omitting colonisation screening would have missed more than a quarter of them. However, no targeted strategy detected more clustered patients per sequenced isolate than random sampling of the same size. Targeted selection therefore did not improve efficiency; it changed which patients could be detected.

Differences in sampling accounted for much, but not all, of the apparent difference between the hospitals. Clustering among clinical, non-screening isolates was similar at both sites, and most of the difference arose from colonisation-screening isolates, which were extensively sequenced in Hannover but largely excluded by Copenhagen’s resistance-based screening policy. Even after adjustment for ICU association, resistance status, and specimen type, clustering remained more common in Hannover during the matched 18-month period (adjusted OR 1.49, 95% CI 1.28-1.73), The difference disappeared only when the full 50-month Copenhagen dataset was analysed, which included more patients and covered a longer period for clusters to accumulate. A real difference in transmission between the hospitals therefore remains possible, but cannot be distinguished from sampling differences and other unmeasured factors by these analyses.

The extent of sequencing mattered just as much as the type of specimen. Sequencing additional isolates from patients already in the dataset also revealed new cluster memberships, since individual patients sometimes carried more than one clone. The 14-16% of patients assigned to a cluster should therefore be regarded as a lower estimate rather than a final measure of the burden of clustering.

The 30-SNP threshold did not capture related isolates equally well across the three species, and the relationship between genomic clustering and ward contact also differed between them. Shared ward contact was most common among clustered patients with *K. pneumoniae*, which is compatible with transmission through gut carriage and with screening-detected carriage contributing to the higher clustering rate in Hannover.^13^ It was least common among patients with *E. coli*, most of whom had no documented ward contact at all; genomic similarity alone therefore does not demonstrate transmission on a particular ward. In *P. aeruginosa*, within-patient diversity often exceeded 30 SNPs (Figure S2C), so related isolates were more likely to be split between clusters and clustering in this species was probably underestimated. Direct ward contacts were also observed across the full range of genetic distances; this pattern is compatible with a shared environmental source, such as contaminated water outlets or sinks, but does not distinguish it from direct patient-to-patient transmission.^18^

This two-centre study was conducted in tertiary-care hospitals in northern Europe with low-to-moderate resistance levels. The sites were chosen to assess the effects of sampling, not to provide a formal comparison between countries or institutions, and site, country, screening policy, and resistance levels could not be separated completely. The findings should be tested in other settings. Genomic clusters do not prove transmission, and the analysis relied on a single-linkage approach with one SNP threshold applied to all three species. The interpretation of genetic distances also depends on within-host diversity, sampling completeness, and the methods used to calculate genomic differences.^19–22^Alternative approaches can identify genomic groups without imposing a single fixed distance threshold.^23^ Inpatient-movement data did not capture contacts in outpatient care, the community, or other hospitals, and neither hospital screened all asymptomatic carriers. Incomplete susceptibility testing, particularly in Copenhagen, might have led to underascertainment of MDR.

In our study, most detected clusters involved patients or organisms that current surveillance strategies do not target, and the number detected depended mainly on how many isolates were sequenced rather than on the selection criteria. Sequencing 25,888 isolates is beyond the capacity of most programmes, so the practical question is which isolates to sequence within a limited budget, and whether that investment prevents infections. Modelling at one of the participating hospitals suggests that integrated genomic surveillance could yield health and economic benefits,^15^ but whether a particular sequencing strategy reduces transmission remains untested. Where capacity is limited, it may be preferable to sequence all isolates from a clearly defined ward, species, or time period than to preselect them by resistance or specimen type. Surveillance programmes should report both the composition and the size of their sequenced datasets. This information is needed to compare programmes and to assess the implications of genomic findings for infection control.

## Data Availability

The complete R and Python code for data processing and statistical analysis is publicly available at https://codebase.helmholtz.cloud/mobabioinf/sampling-effort-clustering. Aggregate study results are provided in the manuscript and supplementary material. Raw sequencing reads, genome assemblies, and individual-level analytical data are not shared as part of this publication.

## Author contributions

S.H. conceived and supervised the study and acquired funding. J.E. performed bioinformatic analyses; R.L.M. contributed to bioinformatic analyses. L.K., K.L.N., and D.St. contributed to data acquisition. J.E. and D.St. performed epidemiological analyses. D.Sch. and F.B.H. supervised clinical microbiological work. J.E. and L.K. led data analysis; L.K. and J.E. directly accessed and verified the underlying data reported in the manuscript; L.K. performed data visualization. L.K., J.E., and S.H. interpreted the data and wrote the manuscript. All authors contributed to revision of the manuscript and approved the final version.

All authors had full access to the data and had final responsibility for the decision to submit.

## Acknowledgment

We gratefully acknowledge Sonja Lekovic, Maria-Louise-Braarup Desauv, Helene Emiliussen, Anna-Lena Hagemann and Astrid Dröge for their dedicated technical assistance in clinical isolate collection, DNA extraction, and library preparation for Illumina sequencing, without which this study would not have been possible. S.H. was funded by the Novo Nordisk Foundation (NNF 18OC0033946), and received funding from the Deutsche Forschungsgemeinschaft (DFG, German Research Foundation) under Germany’s Excellence Strategy - EXC 2155 “RESIST” - Project ID 390874280, within the SFB/TRR-298-SHRI - Project-ID 426335750 and in the SPP 2389 (HA 3299/9-1, AOBJ: 687646),

## Declaration of generative Al and AI-assisted technologies

During preparation of this work, the authors used ChatGPT and Codex (OpenAI; models used: GPT-5.6-S01 and GPT-6-Astra) and Claude and Claude Code (Anthropic; model used: Claude Opus 5) to assist with analysis-code revision and commenting, checking of analysis outputs, and revision of manuscript text. The authors reviewed the code, outputs, and text and take responsibility for the analyses and final manuscript.

