## Supplementary Material for "Sampling composition and sequencing effort in the detection of within-hospital Gram-negative genomic clustering: a prospective observational study at two tertiary-care hospitals"

Prof. Dr. Susanne Häussler,

**Content**


| | |
|---|---|
| Supplementary Methods | 2 |
| Supplementary Results | 4 |
| Supplementary Figures | 6 |
| Supplementary Tables | 8 |
| Supplementary References | 15 |

**Supplementary Methods**

**Study population and observation periods**

The final analysed isolates were collected from Nov 2, 2019, to Dec 29, 2023, in Copenhagen and from March 2, 2021, to Aug 24, 2022, in Hannover. The latter dates defined the common calendar-window analysis. Study size was determined by all eligible isolates available within these collection periods; no formal sample-size calculation was performed. All isolates of the three species identified by the two clinical microbiology laboratories during these periods were eligible; the only selection concerned multiple isolates of the same species from one sample (Methods). Patients were counted once per hospital for overall analyses and once per hospital and species for species-specific population summaries (Table S1).

**DNA extraction and whole-genome sequencing**

DNA was extracted using a modified spin-column protocol described earlier,[1] with lysozyme and proteinase K lysis followed by purification on EZ-96 DNA Filter Plates. Libraries were prepared using Hackflex.[2] Sequencing was performed on Illumina NextSeq 500 instruments with 96 libraries per run or NovaSeq 6000 instruments with 384 libraries per run, generating 150-bp paired-end reads.

**Bioinformatics workflow and quality control**

Raw reads were quality filtered and taxonomically classified. Only isolates assigned to *Escherichia coli*, *Klebsiella pneumoniae* sensu stricto, or *Pseudomonas aeruginosa* were retained; other members of the *K. pneumoniae* species complex were excluded. Sequence types (STs) were assigned using PubMLST schemes. Genomes were assembled with Shovill 1.1.0 using SPAdes,[3,4] assessed with QUAST 5.0.2,[5] and annotated with Prokka 1.14.6.[6] Assemblies with more than 500 contigs or outside the species-specific genome-size ranges were excluded. Accepted ranges were 4.29–6.05 Mb for *E. coli*, 5.00–6.30 Mb for *K. pneumoniae*, and 5.46–7.70 Mb for *P. aeruginosa*. The final dataset contained 25,888 genomes. Workflows were managed with Snakemake 7.0.0[7] in Conda environments.

**Genomic distances and cluster definition**

Genomes were grouped using a gene-content graph with Jaccard similarity of at least 0.80. Pairwise single-nucleotide polymorphism (SNP) distances were calculated within candidate groups using ska.rust 0.3.2 with ambiguous positions filtered.[8] Where multiple estimates existed for an unordered isolate pair, the minimum was retained. Primary single-linkage edges required an eligible direct Jaccard relationship and a distance of 30 SNPs or fewer. Adding the other available relationships at this SNP threshold did not change the final connected-component partition. Recombination was not masked. Split k-mer analysis counts a difference only where the flanking k-mers match exactly, so recombined regions largely drop out of the comparison instead of inflating SNP counts, and the approach is therefore relatively insensitive to recombination.

Components were defined across both hospitals. A local cluster was the species-specific and hospital-specific subset containing at least two unique patients. Patients could belong to multiple clusters but were counted once per hospital for overall proportions. Cluster membership was reassessed after subsampling. For threshold selection only, within-hospital isolate pairs were restricted to sampling dates no more than 60 days apart and distances of 120 SNPs or fewer. The minimum eligible distance was summarised for each unordered patient-pair relationship, including same-patient relationships. These restrictions were not applied to primary cluster definition or single-linkage-chaining assessment.

Components were also reconstructed at thresholds of 20 and 15 SNPs. A separate analysis increased the *P. aeruginosa* threshold to 70 SNPs while retaining 30 SNPs for the other species. Chaining was assessed from all observed pairwise isolate distances within each local cluster, including matrix completeness and maximum within-cluster distances (Tables S4A–B and S7).

**Specimen categories and ICU association**

Specimen terms were harmonised across the laboratory information systems (Table S2). Screening status was determined independently and overrode specimen type. All remaining urine, blood, respiratory, wound, and other categories represented clinical, non-screening isolates; catheter and drain specimens were included in other. At patient level, each category indicator was positive if any isolate belonged to that category, allowing patients to contribute to several categories through different isolates. Intensive care unit (ICU) association indicated at least one isolate sampled on an ICU ward and did not capture every previous ICU admission.

### Multidrug resistance

Antimicrobial susceptibility testing used disk diffusion in Copenhagen, supplemented by gradient diffusion where available, and minimum inhibitory concentration determination in Hannover. We used the interpretations reported under the European Committee on Antimicrobial Susceptibility Testing (EUCAST) breakpoint versions applicable at the time of testing, without retrospective harmonisation to a single version. Multidrug resistance (MDR) was defined as resistance (category R) to at least one tested agent in three or more antimicrobial categories, using species-specific panels based on the Magiorakos framework.[9] Category I results did not count as resistant. Panels prioritised agents commonly tested at both hospitals and did not cover the full original framework.

Missing or untested results did not contribute to resistant-category counts, and no minimum number of tested categories was required. Fewer than three categories had an interpretable result for 113 of 13,248 Copenhagen isolates and one of 12,640 Hannover isolates. These isolates remained classified as non-MDR. One Copenhagen isolate was retained as MDR on the basis of a laboratory-reported OXA-48 carbapenemase finding despite not meeting the phenotypic three-category criterion; carbapenemase information was not systematically available for the remaining isolates. Non-MDR did not imply susceptibility to every agent.

For *E. coli*, the assessed categories were aminoglycosides (gentamicin or tobramycin), antipseudomonal penicillins with inhibitors (piperacillin–tazobactam), carbapenems (imipenem or meropenem), second-generation cephalosporins (cefuroxime), extended-spectrum cephalosporins (cefotaxime in Hannover or ceftriaxone in Copenhagen, or ceftazidime), fluoroquinolones (ciprofloxacin), folate-pathway inhibitors (trimethoprim–sulfamethoxazole), glycylcyclines (tigecycline), penicillins (ampicillin), penicillins with inhibitors (ampicillin–sulbactam in Hannover or amoxicillin–clavulanate in Copenhagen), and phosphonic acids (fosfomycin). The same categories were used for *K. pneumoniae* except ampicillin. For *P. aeruginosa*, the categories were aminoglycosides (tobramycin), antipseudomonal penicillins with inhibitors (piperacillin–tazobactam), carbapenems (imipenem or meropenem), antipseudomonal cephalosporins (ceftazidime), fluoroquinolones (ciprofloxacin), and phosphonic acids (fosfomycin).

### Diversity and regression analyses

ST richness was the number of observed typing categories. Untypeable isolates formed one additional category in the primary analysis and were excluded in a sensitivity analysis. Gini–Simpson diversity was calculated as $1 - \Sigma p_i^2$, where $p_i$ was the isolate-level frequency of category i. Patient-based rarefaction retained all isolates from selected patients and used 1000 random samples without replacement. Curves show mean richness with 2.5th–97.5th percentile bands; matched comparisons used the smaller hospital-specific patient count for each species (Figure S1; Table S3).

Patient-level binomial generalised linear models with a logit link used membership in any local genomic cluster as the outcome. Covariates were hospital, any ICU-associated isolate, any MDR isolate, and indicators for urine, blood, respiratory, screening, wound, and other specimens. Odds ratios (ORs) compared Hannover with Copenhagen or presence with absence of an indicator. Model-based 95% confidence intervals (CIs) and two-sided Wald p values were reported.

The common-window analysis included all isolates collected at both hospitals from March 2, 2021, to Aug 24, 2022, comprising 3229 Copenhagen patients and 5450 Hannover patients. Cluster membership and patient-level covariates were reconstructed using only isolates collected within this window. This analysis did not involve random subsampling.

Patient-number matching was a separate analysis based on the full Copenhagen cohort: in each of 1000 draws, 5450 of the 8262 Copenhagen patients were sampled without replacement and compared with all 5450 Hannover patients, retaining all isolates from selected patients. The original observation periods were retained. Components and covariates were reconstructed; removing Copenhagen isolates could therefore also affect Hannover cluster membership. Reported ORs and model-based CI limits are medians across draws. Additional 2.5th–97.5th percentile ranges describe variation in OR point estimates across draws, not model-based CIs. Model-specification analyses sequentially varied ICU, MDR, and specimen indicators; omitting the screening indicator did not exclude screening isolates (Tables S6A–B).

### Sequencing effort and targeted strategies

Random sampling without replacement varied the numbers of isolates or patients retained; patient sampling retained all isolates from selected patients. Depth analyses started with one randomly selected isolate per patient and added increasing random fractions of the remaining isolates. The other hospital's isolates remained a fixed genomic background. Components were reconstructed after selection, and local detection required at least two patients at the hospital being evaluated. Figure 3 reports means across 500 draws and 10th–90th percentile bands in panels A and B. Qualifying fragments of a full-data component were counted separately in panel A. Complete-data endpoints were evaluated directly, without extrapolation beyond the observed sampling range.

Hannover strategies comprised clinical-only, screening-only, ICU-associated, urine, respiratory, blood-culture, MDR-only, and all clinical plus MDR screening isolates. Specimen-type strategies included clinical, non-screening isolates only. Each strategy was compared with 500 random subsets of the same number of Hannover isolates, retaining Copenhagen isolates as a fixed background. Figure 5 reports random-sampling medians and 2.5th–97.5th percentiles. Unlike Figure 3A, each full-data reference cluster was counted at most once if any surviving fragment contained at least two Hannover patients.

Screening-policy analyses retained all clinical isolates and either omitted all screening isolates or retained only MDR screening isolates. The latter was not an exact reconstruction of Copenhagen's carbapenem-resistance-based policy. Clinical-only and screening-only detected-patient sets were compared by intersection and set differences; patients detected in the full dataset but in neither restricted analysis required the combined dataset.

**Inpatient-movement contacts**

Inpatient histories were obtained from Hannover Medical School's Enterprise Clinical Research Data Warehouse and Rigshospitalet's forskerplatform. The same contact definitions were used at both hospitals. For each patient pair within a cluster, ward stays beginning on or after the later of the two patients' first cluster-isolate sampling dates were excluded. Direct contact required overlapping recorded stays on the same ward. For indirect contact, the earlier-detected patient's stay end was extended by 14 days and the later-detected patient's stay start was moved 14 days earlier; overlap of the adjusted same-ward intervals defined an indirect contact when no direct contact was present. For tied first sampling dates, both stay ends were extended. Direct contacts took precedence over indirect contacts. These rules did not establish transmission direction.

Species-specific analyses used patient–species–cluster assignments. Overall hospital estimates counted each patient once and classified them as linked if any assignment had a documented direct or indirect contact with a cluster co-member. Unavailable movement data were retained as no documented link in the conservative analysis. Complete-case analyses excluded unavailable assignments; unique patients were excluded only if data were unavailable for all their assignments. Local clusters were counted once and considered supported if any documented contact was present. Proportions were reported with Wilson 95% CIs (Tables S5A–B).

Nearest-neighbour analyses used the minimum observed distance to an isolate from another patient in the same local cluster. Where several co-members were tied, direct contact took precedence over indirect contact and then no identifiable contact. These estimates differ from contact with any cluster co-member. Table S5C includes nearest distances of 30 SNPs or fewer; larger distances are displayed separately in Figure 1D–F.

**Ward-contact permutation analysis**

As a separate supportive analysis, proxy contacts required isolates from two patients to have been sampled on the same ward less than 14 days apart. Cluster labels were permuted across patient–species–cluster assignment rows within each hospital and species, preserving the contact network and the number of rows carrying each cluster label. Patients with several assignments contributed several rows; unique patient membership within each permuted cluster was not enforced. The empirical one-sided p value was (1 + number of permuted proportions at least as large as observed)/(1000 + 1). This sampling-event proxy did not replace inpatient-movement histories or establish a transmission mechanism (Table S5D).

**Software and reproducibility**

Analyses used Python 3.13.3, pandas 2.3.0, NumPy 2.2.6, and statsmodels 0.14.5. Figures were generated with Altair 6.2.2 and vl-convert-python 1.9.0.post1. Fixed random seeds were used for simulations and permutations, as specified in the published R and Python code. Code availability is described in the main manuscript.

**Supplementary Results**

**Specimen composition and clinical representation**

Urine and blood isolates accounted for 54.7% and 6.6% of Copenhagen isolates and 33.7% and 3.2% of Hannover isolates. Respiratory specimens represented 11.3% and 20.2%, respectively. Copenhagen contributed 38 screening isolates and Hannover 2796; 1013 Hannover screening isolates (36.2%) were non-MDR. At isolate level, 12.7% of Copenhagen isolates and 19.6% of Hannover isolates belonged to a local cluster (Figure 2; Tables S1–2). Patient-level clustering was 13.8% in Copenhagen and 15.1% in Hannover among patients with urine isolates and 32.0% and 25.8%, respectively, among patients with screening isolates. These category-specific proportions did not constitute a comparison of mutually exclusive patient groups.

MDR isolates were present in 28.7% of clustered and 21.0% of non-clustered patients in Copenhagen, and 50.0% and 40.0% in Hannover. Among clustered patients without an ICU-associated isolate, 73.2% in Copenhagen and 53.7% in Hannover had no MDR isolate. Among *P. aeruginosa* local clusters, 90.6% in Copenhagen and 60.3% in Hannover contained no MDR isolate. Clustering was more frequent among patients with an ICU-associated isolate than among those without one (21.2% vs 13.3% in Copenhagen; 23.7% vs 14.5% in Hannover).

Among full-data clustered patients, 1158 of 1161 in Copenhagen (99.7%) and 717 of 900 in Hannover (79.7%) had a clinical, non-screening isolate within a valid full-data component. Having a clinical isolate in the full dataset was distinct from being detectable after restricting the analysis to clinical isolates: in Hannover, the clinical-only analysis detected 660 clustered patients (73.3%).

**Additional epidemiological and threshold findings**

Among *E. coli* patient–species–cluster assignments, nearest-neighbour contact proportions were 35.2% at 0–5 SNPs, 6.7% at 6–15 SNPs, and 4.8% at 16–30 SNPs when both hospitals were pooled (Table S5C). These percentages referred to contact with a nearest genomic co-member, rather than any co-member. The full species-specific contact proportions and confidence intervals are provided in Table S5A.

The largest observed within-cluster pairwise distance was 1557 SNPs. Increasing the *P. aeruginosa* threshold to 70 SNPs increased the number of local clusters from 64 to 86 in Copenhagen and from 73 to 104 in Hannover, alongside the increased clustered-patient counts reported in the main Results (Table S7). This sensitivity analysis did not establish 70 SNPs as a validated transmission cutoff.

**Regression and standardisation**

In the full adjusted model, ORs were 1.35 (95% CI 1.17–1.55) for ICU association, 1.21 (1.08–1.35) for MDR, 1.56 (1.37–1.77) for urine, 1.61 (1.36–1.91) for blood, 1.58 (1.36–1.85) for respiratory specimens, 2.22 (1.84–2.67) for screening, 1.85 (1.60–2.13) for wound specimens, and 1.55 (1.35–1.79) for other specimens. Each specimen indicator compared presence with absence of that category. The full-cohort hospital p value was below 0.0001 before adjustment and 0.49 after adjustment.

In the common calendar window, 305 of 3229 Copenhagen patients and 894 of 5450 Hannover patients clustered, giving a crude hospital OR of 1.88 (95% CI 1.64–2.16). Patient-number matching yielded a median crude OR of 1.53 (median model-based 95% CI 1.37–1.70; empirical 2.5th–97.5th percentile range 1.42–1.66) and a median adjusted OR of 1.23 (median model-based 95% CI 1.08–1.39; empirical percentile range 1.14–1.33; Table S6A). The full-cohort and common-window model-specification analyses are reported in Table S6B.

**Sequencing effort and detection per isolate**

At 25% isolate sampling, 3312 Copenhagen isolates yielded a mean of 73.6 detected local clusters and 3160 Hannover isolates yielded 85.7. The corresponding patient-based subsamples contained 2066 and 1362 patients and yielded mean clustered-patient proportions of 6.4% and 7.9%, respectively (Figure 3).

Across the complete datasets, 900 clustered patients were detected among 12,640 isolates in Hannover (71.2 per 1000 isolates), compared with 1161 among 13,248 isolates in Copenhagen (87.6 per 1000). Patients contributed a mean of 2.32 and 1.60 isolates, respectively. Detection per 1000 isolates equals the proportion of patients detected as clustered divided by the mean number of isolates per patient, multiplied by 1000. These descriptive ratios therefore depend on both patient-level detection and repeated sampling and are not an independent measure of transmission intensity or programme efficiency.

**Targeted strategies and screening contribution**

In Hannover, respiratory-only sequencing retained 2551 isolates and detected 96 clustered patients, compared with a random-sampling median of 173 (2.5th–97.5th percentile range 149–201). ICU-only sequencing retained 3124 isolates and detected 154 patients, compared with 219 (188–246); urine-only sequencing retained 4258 isolates and detected 274 patients, compared with 307 (275–345); and MDR-only sequencing retained 5138 isolates and detected 337 patients, compared with 376 (344–412). Retaining all clinical isolates and MDR screening isolates included 11,627 isolates and detected 791 patients, compared with 836 (814–853). Clinical-only sequencing retained 9844 isolates and detected 660 patients, compared with 717 (688–744). Blood-only and screening-only results are reported in the main Results (Figure 5A). Percentile endpoints are rounded to whole patients.

Clinical-only and screening-only analyses detected 660 and 185 clustered patients, respectively; their intersection contained 39 patients and their union 806. A further 94 patients were detected only when the isolate sets were analysed together. Of the 240 patients missed without screening, 40.0% had no MDR isolate; among the 109 patients missed under MDR-selected screening, 82.6% had no MDR isolate. Detection through screening alone did not establish that a patient had no clinical specimen, because the available clinical isolates might be insufficient to establish local cluster membership (Figure 5B–C).

The Hannover ST86 cluster spanned 310 days across 11 relative calendar months. Its five clinical isolates included four blood isolates from three patients, supplementing the 169 screening isolates described in the main Results (Figure 4B). The analysis did not reconstruct detection of this individual cluster under Copenhagen's exact carbapenem-resistance-based screening policy.

### Supplementary Figures
### Figure S1

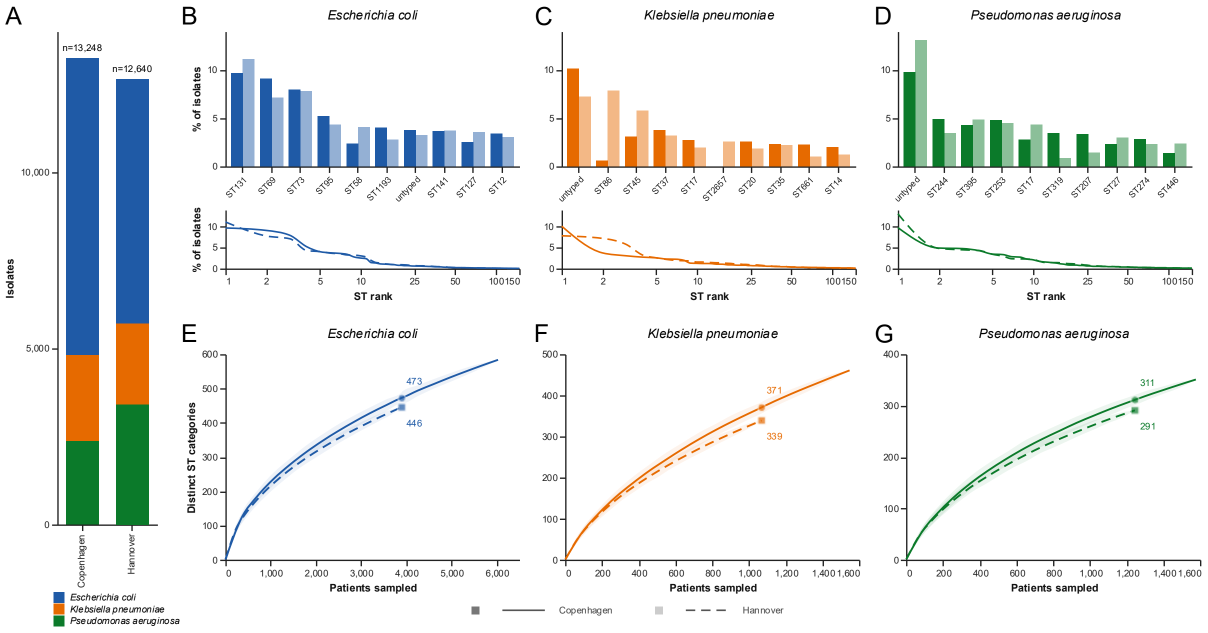


**Figure S1. Composition and diversity of sequence types at each hospital.** (A) Numbers of sequenced isolates by species and hospital. (B–D) The ten most frequent typing categories for *Escherichia coli*, *Klebsiella pneumoniae*, and *Pseudomonas aeruginosa*, respectively, with rank-frequency profiles up to rank 150 on a logarithmic rank axis. Darker bars represent Copenhagen and lighter bars Hannover. (E–G) Patient-based rarefaction curves for the corresponding species, retaining all isolates from sampled patients. Lines show mean numbers of distinct typing categories; shaded bands show the 2.5th–97.5th percentiles of 1000 random samples. Solid and dashed lines represent Copenhagen and Hannover, respectively, in the rank-frequency profiles and rarefaction curves. Marked estimates compare hospitals at the smaller patient count within each species. Untypeable isolates were retained as one additional category. ST=sequence type.

**Figure S2**

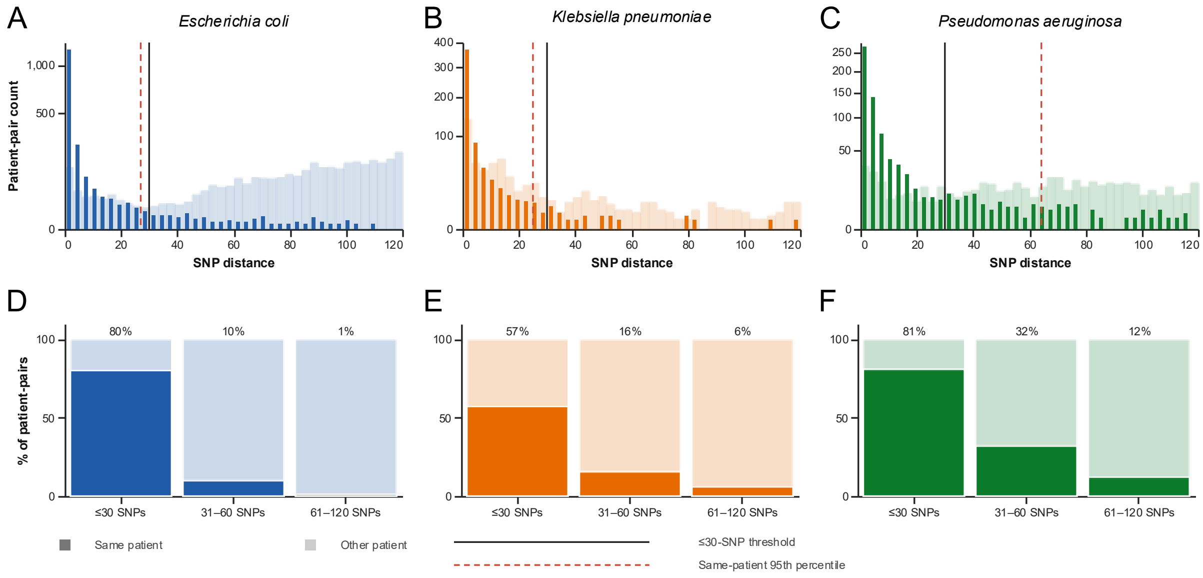


**Figure S2. Within-patient genomic diversity and threshold selection.** (A–C) Distributions of minimum eligible SNP distances for same-patient and between-patient relationships in *Escherichia coli*, *Klebsiella pneumoniae*, and *Pseudomonas aeruginosa*, respectively. Isolate pairs from the same hospital were included if sampling dates were no more than 60 days apart and genomic distances were 120 SNPs or fewer. The minimum eligible distance was summarised for each patient-pair relationship within each species. Darker shading indicates same-patient relationships and lighter shading between-patient relationships. Solid black lines mark 30 SNPs; red dashed lines mark the same-patient 95th percentiles: 27 SNPs for *E. coli*, 25 for *K. pneumoniae*, and 64 for *P. aeruginosa*. (D–F) Same-patient and between-patient proportions in the corresponding species within the ≤30, 31–60, and 61–120 SNP distance groups. Percentages above the bars show the same-patient fraction. The 60-day and 120-SNP restrictions were not applied to primary cluster definition or single-linkage-chaining assessment. SNP=single-nucleotide polymorphism.

### Supplementary Tables

#### Table S1. Cohort and sampling characteristics

| | Copenhagen | Hannover |
|---|---|---|
| First sampling date | 2019-11-02 | 2021-03-02 |
| Last sampling date | 2023-12-29 | 2022-08-24 |
| Isolates (n) | 13,248 | 12,640 |
| Patients (n) | 8262 | 5450 |
| Screening isolates (n) | 38 | 2796 |
| Screening isolates (%) | 0.29 | 22.12 |
| ICU isolates (n) | 1241 | 3124 |
| ICU isolates (%) | 9.37 | 24.72 |
| MDR isolates (n) | 2932 | 5138 |
| MDR isolates (%) | 22.13 | 40.65 |
| *E. coli* (n) | 8438 | 6945 |
| *K. pneumoniae* (n) | 2448 | 2286 |
| *P. aeruginosa* (n) | 2362 | 3409 |

Patients were counted once per hospital. MDR classification used resistance to at least one tested agent in three or more antimicrobial categories, with one laboratory-reported exception; testing coverage and classification details are described in the Supplementary Methods. ICU=intensive care unit. MDR=multidrug-resistant.

#### Table S2. Harmonisation of specimen categories between hospitals

| Raw specimen term | Specimen type | Final analytical category | Isolates (n) |
|---|---|---|---|
| Urin (midtstråle) | Urine | Urine | 4630 |
| Mid-stream urine sample | Urine | Urine | 2571 |
| Urin fra KAD | Urine | Urine | 1364 |
| Urine specimen obtained via indwelling urinary catheter | Urine | Urine | 1321 |
| Urin fra reservoir (Bricker) | Urine | Urine | 519 |
| Urin | Urine | Urine | 294 |
| Urin (nefrostomikateter) | Urine | Urine | 226 |
| Urin (suprapubisk punktur) | Urine | Urine | 174 |
| Suprapubic urine specimen | Urine | Urine | 124 |
| Renal pelvis fluid sample | Urine | Urine | 99 |
| Urine specimen obtained from pediatric urine collection bag | Urine | Urine | 92 |
| Urine specimen from urinary conduit | Urine | Urine | 47 |
| Urin fra J-J kateter | Urine | Urine | 40 |
| Urine specimen | Urine | Urine | 3 |
| First stream urine sample | Urine | Urine | 1 |
| Urin (kateter) | Urine | Urine | 1 |
| Urin (suprapubisk/topkateter) | Urine | Urine | 1 |
| Blod fra perifer vene (kolbe) | Blood | Blood | 607 |
| Blood specimen in aerobic blood culture bottle | Blood | Blood | 272 |
| Blod fra kateter (kolbe) | Blood | Blood | 245 |
| Blood specimen in anaerobic blood culture bottle | Blood | Blood | 133 |
| Blod fra arterie (kolbe) | Blood | Blood | 19 |
| Blod (bloddyrkningskolbe) | Blood | Blood | 3 |
| Sputum specimen | Respiratory | Respiratory | 889 |
| Trachealsekret | Respiratory | Respiratory | 779 |
| Ekspektorat | Respiratory | Respiratory | 642 |
| Specimen from trachea obtained by aspiration | Respiratory | Respiratory | 520 |
| Swab specimen from oropharynx | Respiratory | Respiratory | 446 |
| Bronchoalveolar lavage fluid sample | Respiratory | Respiratory | 404 |
| Bronchial fluid sample | Respiratory | Respiratory | 169 |
| BAL | Respiratory | Respiratory | 76 |
| Specimen from lung obtained by bronchial washing procedure | Respiratory | Respiratory | 58 |
| Sinus washings | Respiratory | Respiratory | 29 |
| Swab of internal nose | Respiratory | Respiratory | 23 |
| Pharyngeal washings | Respiratory | Respiratory | 6 |
| Nasopharyngeal swab | Respiratory | Respiratory | 3 |
| Oral swab | Respiratory | Respiratory | 3 |
| Swab from tongue | Respiratory | Respiratory | 1 |
| Rectal swab | Other | Screening | 2099 |
| Swab specimen from oropharynx | Respiratory | Screening | 192 |
| Stool specimen | Other | Screening | 154 |
| Nasopharyngeal swab | Respiratory | Screening | 142 |
| Swab of groin | Other | Screening | 82 |
| Anal swab | Other | Screening | 45 |
| Podning | Other | Screening | 36 |
| Specimen from trachea obtained by aspiration | Respiratory | Screening | 31 |
| Mid-stream urine sample | Urine | Screening | 18 |
| Swab of axilla | Other | Screening | 7 |
| Urine specimen obtained via indwelling urinary catheter | Urine | Screening | 6 |
| Swab from superficial wound | Wound | Screening | 5 |

| Raw specimen term | Specimen type | Final analytical category | Isolates (n) |
|---|---|---|---|
| Sputum specimen | Respiratory | Screening | 4 |
| Wound swab | Wound | Screening | 4 |
| Swab | Other | Screening | 3 |
| Podning sår | Wound | Screening | 2 |
| Swab of internal nose | Respiratory | Screening | 2 |
| Drainage fluid sample | Other | Screening | 1 |
| Specimen | Other | Screening | 1 |
| Swab from surgical wound | Wound | Wound | 589 |
| Swab from superficial wound | Wound | Wound | 548 |
| Podning sår | Wound | Wound | 416 |
| Podning cicatrice | Wound | Wound | 323 |
| Swab from deep wound | Wound | Wound | 300 |
| Pus (i glas) | Wound | Wound | 259 |
| Podning brandsår | Wound | Wound | 223 |
| Wound swab | Wound | Wound | 144 |
| Podning absces | Wound | Wound | 75 |
| Specimen from abscess obtained by aspiration | Wound | Wound | 70 |
| Podning decubitus | Wound | Wound | 54 |
| Podning pus | Wound | Wound | 22 |
| Podning - væv | Wound | Wound | 5 |
| Hæmatom | Wound | Wound | 4 |
| Podning indstikssted | Wound | Wound | 3 |
| Absces (aspirat) | Wound | Wound | 2 |
| Podning sekret | Wound | Wound | 2 |
| Podning bidsår | Wound | Wound | 1 |
| Podning | Other | Other | 858 |
| Drænvæske | Other | Other | 289 |
| Væv | Other | Other | 231 |
| Drainage fluid sample | Other | Other | 207 |
| Kateterspids | Other | Other | 200 |
| Bile specimen | Other | Other | 151 |
| Swab | Other | Other | 91 |
| Aspirat (opsug) | Other | Other | 88 |
| Væske | Other | Other | 83 |
| Ansamling | Other | Other | 79 |
| Cervical swab | Other | Other | 76 |
| Vaginal swab | Other | Other | 72 |
| Swab of groin | Other | Other | 67 |
| Specimen obtained by aspiration | Other | Other | 47 |
| Sekret | Other | Other | 45 |
| Drænspids | Other | Other | 44 |
| Kammebiopsi | Other | Other | 42 |
| Specimen from prosthetic joint | Other | Other | 41 |
| Pleuravæske | Other | Other | 40 |
| Seminal fluid specimen | Other | Other | 33 |
| Ascitic fluid sample | Other | Other | 30 |
| Biopsi | Other | Other | 26 |
| Tissue specimen | Other | Other | 26 |
| Ascites/peritonealvæske | Other | Other | 22 |
| Urethral swab | Other | Other | 22 |
| Conjunctival swab | Other | Other | 20 |
| Implantationsmateriale | Other | Other | 20 |
| Stent | Other | Other | 17 |
| Ear swab sample | Other | Other | 16 |
| Specimen | Other | Other | 16 |
| Fæces | Other | Other | 14 |
| Knoglevæv | Other | Other | 14 |
| Implantat/protese sonikering | Other | Other | 13 |
| Serom | Other | Other | 12 |
| Swab of axilla | Other | Other | 12 |
| Afskrab | Other | Other | 10 |
| Ledvæske | Other | Other | 9 |
| Lymph node sample | Other | Other | 9 |
| Vascular catheter submitted as specimen | Other | Other | 9 |
| Peritonealdialysevæske (kolbe) | Other | Other | 8 |
| Sædprøve | Other | Other | 7 |
| Biopsy sample | Other | Other | 6 |
| Pleural fluid specimen | Other | Other | 6 |
| Spinalvæske | Other | Other | 6 |
| Ammemælk | Other | Other | 5 |
| Cystevæske | Other | Other | 5 |
| Catheter submitted as specimen | Other | Other | 4 |
| Kultur | Other | Other | 4 |
| Contact lens submitted as specimen | Other | Other | 3 |
| Knoglemarv | Other | Other | 3 |

| Raw specimen term | Specimen type | Final analytical category | Isolates (n) |
|---|---|---|---|
| Kontaktlinse | Other | Other | 3 |
| Amnion cytologic material | Other | Other | 2 |
| Anal swab | Other | Other | 2 |
| Drain tip submitted as specimen | Other | Other | 2 |
| Peritoneal dialysis fluid specimen | Other | Other | 2 |
| Swab of eye | Other | Other | 2 |
| Ascitesvæske (kolbe) | Other | Other | 1 |
| Fostervand | Other | Other | 1 |
| Hemodialysis catheter submitted as specimen | Other | Other | 1 |
| Intracranial ventricular catheter submitted as specimen | Other | Other | 1 |
| Joint fluid specimen | Other | Other | 1 |
| Pericardievæske | Other | Other | 1 |
| Prostatic fluid sample | Other | Other | 1 |
| Rectal swab | Other | Other | 1 |
| Øjelinse | Other | Other | 1 |

Raw specimen terms were retrieved from the hospitals' laboratory information systems. Hannover terms had been mapped to SNOMED CT specimen concepts before harmonisation, whereas Copenhagen terms retained the local Danish terminology. Terms were mapped to common specimen types. Screening status was determined independently and overrode the specimen type in the final analytical variable. Screening specimens were therefore counted only as Screening and not additionally under urine, respiratory, wound, or other categories. For non-screening isolates, the final analytical category corresponds to the specimen type. Isolate counts are pooled across both hospitals. SNOMED CT=Systematized Nomenclature of Medicine Clinical Terms.

**Table S3. Sequence-type diversity and sensitivity to the handling of untypeable isolates**

| | Copenhagen | | | Hannover | | |
|---|---|---|---|---|---|---|
| | *E. coli* | *K. pneumoniae* | *P. aeruginosa* | *E. coli* | *K. pneumoniae* | *P. aeruginosa* |
| Isolates excluding untypeable ST (n) | 8120 | 2200 | 2131 | 6719 | 2121 | 2963 |
| Isolates retaining untypeable ST (n) | 8438 | 2448 | 2362 | 6945 | 2286 | 3409 |
| Patients excluding untypeable ST (n) | 5819 | 1385 | 1428 | 3785 | 981 | 1112 |
| Patients retaining untypeable ST (n) | 6028 | 1548 | 1577 | 3898 | 1069 | 1245 |
| Richness excluding untypeable ST | 583 | 460 | 350 | 445 | 338 | 290 |
| Richness retaining untypeable ST | 584 | 461 | 351 | 446 | 339 | 291 |
| Gini–Simpson diversity excluding untypeable ST | 0.96 | 0.99 | 0.98 | 0.96 | 0.98 | 0.98 |
| Gini–Simpson diversity retaining untypeable ST | 0.96 | 0.98 | 0.98 | 0.96 | 0.98 | 0.97 |

Sequence-type richness and Gini–Simpson diversity were calculated from all isolates within each hospital and species. In the primary analysis, untypeable isolates were retained as one additional typing category; in the sensitivity analysis, they were excluded completely. Patients were counted once per hospital and species within each analysis. Gini–Simpson diversity was calculated as $1 - \Sigma p_i^2$, where $p_i$ is the proportion of isolates belonging to typing category i. ST=sequence type.

**Table S4. Robustness of local-cluster definition**
**Table S4A. SNP-threshold sensitivity**

| | Copenhagen | | | Hannover | | |
|---|---|---|---|---|---|---|
| Patients total (n) | | | 8,262 | | | 5,450 |
| Isolates total (n) | | | 13,248 | | | 12,640 |
| **Cutoff** | **≤30 SNPs** | **≤20 SNPs** | **≤15 SNPs** | **≤30 SNPs** | **≤20 SNPs** | **≤15 SNPs** |
| Clustered patients (n) | 1161 | 922 | 755 | 900 | 738 | 637 |
| Clustered patients (%) | 14.05 | 11.16 | 9.14 | 16.51 | 13.54 | 11.69 |
| Clustered isolates (n) | 1679 | 1352 | 1133 | 2479 | 2126 | 1892 |
| Clustered isolates (%) | 12.67 | 10.21 | 8.55 | 19.61 | 16.82 | 14.97 |
| Local clusters (n) | 388 | 326 | 274 | 340 | 280 | 242 |
| Mean isolates per cluster | 4.33 | 4.15 | 4.14 | 7.29 | 7.59 | 7.82 |
| Median isolates per cluster | 3 | 3 | 3 | 4 | 4 | 4 |

Global genomic components were reconstructed by single-linkage clustering separately at each SNP threshold. A local cluster was defined as the hospital-specific and species-specific subset of a global component containing isolates from at least two patients. Patients were counted once per hospital if they belonged to at least one local cluster, whereas cluster size was calculated from the number of isolates. SNP=single-nucleotide polymorphism.

**Table S4B. Single-linkage chaining**

| | Copenhagen | | | Hannover | | |
|---|---|---|---|---|---|---|
| | *E. coli* | *K. pneumoniae* | *P. aeruginosa* | *E. coli* | *K. pneumoniae* | *P. aeruginosa* |
| Local clusters (n) | 252 | 72 | 64 | 190 | 77 | 73 |
| Clusters with all pairs ≤30 SNPs (n) | 214 | 55 | 41 | 155 | 66 | 43 |
| Clusters with all pairs ≤30 SNPs (%) | 84.92 | 76.39 | 64.06 | 81.58 | 85.71 | 58.90 |
| Clusters with at least one pair >30 SNPs (n) | 38 | 17 | 23 | 35 | 11 | 30 |
| Clusters with at least one pair >30 SNPs (%) | 15.08 | 23.61 | 35.94 | 18.42 | 14.29 | 41.10 |
| Clusters with at least one pair >120 SNPs (n) | 10 | 2 | 7 | 7 | 0 | 7 |
| Clusters with at least one pair >120 SNPs (%) | 3.97 | 2.78 | 10.94 | 3.68 | 0.00 | 9.59 |
| Maximum within-cluster SNP distance (median [IQR]) | 17.5 [8–26] | 18.5 [8.5–30] | 26.5 [17.25–47] | 17 [7.25–28] | 11 [5–21] | 26 [16–41] |

The analysis includes local clusters defined using the primary threshold of 30 SNPs or fewer. The maximum within-cluster distance was the largest pairwise isolate distance observed within each local cluster. All pairwise distance matrices were complete. Clusters containing at least one pair separated by more than 30 SNPs arose through single-linkage chaining; clusters containing a pair separated by more than 120 SNPs form a subset of this group. IQR=interquartile range. SNP=single-nucleotide polymorphism.

**Table S5. Epidemiological support for genomic clustering**

**Table S5A. Patient-level contact estimates and missing-data sensitivity**

| | Copenhagen | | | | | Hannover | | | | |
|---|---|---|---|---|---|---|---|---|---|---|
| | **Patient–species–cluster assignments** | **Unique patients** | **Patient–species–cluster assignments** | **Patient–species–cluster assignments** | **Patient–species–cluster assignments** | **Patient–species–cluster assignments** | **Unique patients** | **Patient–species–cluster assignments** | **Patient–species–cluster assignments** | **Patient–species–cluster assignments** |
| | **All species** | **All species** | ***E. coli*** | ***K. pneumoniae*** | ***P. aeruginosa*** | **All species** | **All species** | ***E. coli*** | ***K. pneumoniae*** | ***P. aeruginosa*** |
| Without movement data (n) | 57 | 57 | *39* | *7* | *11* | 129 | 125 | *78* | *18* | *33* |
| Linked, conservative analysis (n) | 249 | 243 | 102 | 81 | 66 | 368 | 335 | 147 | 145 | 76 |
| Denominator, conservative analysis | 1195 | 1161 | 732 | 225 | 238 | 967 | 900 | 515 | 243 | 209 |
| Linked, conservative analysis (%) | 20.84 | 20.93 | 13.93 | 36.00 | 27.73 | 38.06 | 37.22 | 28.54 | 59.67 | 36.36 |
| Lower 95% CI, conservative analysis | 18.63 | 18.69 | 11.61 | 30.01 | 22.43 | 35.05 | 34.12 | 24.81 | 53.40 | 30.14 |
| Upper 95% CI, conservative analysis | 23.23 | 23.36 | 16.63 | 42.46 | 33.74 | 41.16 | 40.43 | 32.59 | 65.64 | 43.08 |
| Linked, complete-case analysis (n) | 249 | 243 | 102 | 81 | 66 | 368 | 335 | 147 | 145 | 76 |
| Denominator, complete-case analysis | 1138 | 1104 | 693 | 218 | 227 | 838 | 775 | 437 | 225 | 176 |
| Linked, complete-case analysis (%) | 21.88 | 22.01 | 14.72 | 37.16 | 29.07 | 43.91 | 43.23 | 33.64 | 64.44 | 43.18 |
| Lower 95% CI, complete-case analysis | 19.58 | 19.67 | 12.28 | 31.02 | 23.55 | 40.59 | 39.78 | 29.37 | 58.00 | 36.09 |
| Upper 95% CI, complete-case analysis | 24.37 | 24.55 | 17.55 | 43.74 | 35.29 | 47.29 | 46.74 | 38.19 | 70.41 | 50.57 |

A documented ward contact comprised a direct or indirect link to another patient in the same genomic cluster. Patients contributing to more than one species or cluster were counted once per patient–species–cluster assignment in the assignment-level analyses and once across all assignments in the unique-patient analyses. The conservative analysis retained assignments without available movement data and classified them as having no documented link. The complete-case analysis excluded assignments without movement data; unique patients were excluded only when movement data were unavailable for all their assignments. Proportions are presented with Wilson 95% CIs. CI=confidence interval.

**Table S5B. Local clusters with at least one documented ward contact**

| | Copenhagen | | | | Hannover | | | |
|---|---|---|---|---|---|---|---|---|
| | All species | *E. coli* | *K. pneumoniae* | *P. aeruginosa* | All species | *E. coli* | *K. pneumoniae* | *P. aeruginosa* |
| Local clusters (n) | 388 | 252 | 72 | 64 | 340 | 190 | 77 | 73 |
| Clusters with at least one documented ward contact (n) | 92 | 43 | 28 | 21 | 131 | 58 | 44 | 29 |
| Clusters with at least one documented ward contact (% of total) | 23.71 | 17.06 | 38.89 | 32.81 | 38.53 | 30.53 | 57.14 | 39.73 |
| Lower 95% CI | 19.75 | 12.92 | 28.47 | 22.57 | 33.51 | 24.42 | 46.01 | 29.29 |
| Upper 95% CI | 28.19 | 22.19 | 50.44 | 45.00 | 43.80 | 37.41 | 67.60 | 51.19 |

Each local cluster was counted once and was classified as having epidemiological support when at least one patient had a documented direct or indirect ward contact with another member of the same cluster. Assignments without available movement data were treated as having no documented link. Proportions are presented with Wilson 95% CIs. CI=confidence interval.

**Table S5C. Epidemiological links by distance to the nearest genomic co-member**

| | | Nearest SNP distance | Assignments (n) | Linked assignments (n) | Linked assignments (%) | Lower 95% CI | Upper 95% CI |
|---|---|---|---|---|---|---|---|
| **Copenhagen** | ***E. coli*** | 0–5 SNPs | 208 | 41 | 19.71 | 14.88 | 25.65 |
| **Copenhagen** | ***E. coli*** | 6–15 SNPs | 275 | 15 | 5.45 | 3.33 | 8.80 |
| **Copenhagen** | ***E. coli*** | 16–30 SNPs | 249 | 10 | 4.02 | 2.20 | 7.23 |
| **Copenhagen** | ***K. pneumoniae*** | 0–5 SNPs | 87 | 41 | 47.13 | 36.98 | 57.51 |
| **Copenhagen** | ***K. pneumoniae*** | 6–15 SNPs | 89 | 16 | 17.98 | 11.38 | 27.23 |
| **Copenhagen** | ***K. pneumoniae*** | 16–30 SNPs | 49 | 2 | 4.08 | 1.13 | 13.71 |
| **Copenhagen** | ***P. aeruginosa*** | 0–5 SNPs | 68 | 17 | 25.00 | 16.24 | 36.44 |
| **Copenhagen** | ***P. aeruginosa*** | 6–15 SNPs | 80 | 16 | 20.00 | 12.70 | 30.05 |
| **Copenhagen** | ***P. aeruginosa*** | 16–30 SNPs | 84 | 3 | 3.57 | 1.22 | 9.98 |
| **Hannover** | ***E. coli*** | 0–5 SNPs | 212 | 107 | 50.47 | 43.79 | 57.13 |
| **Hannover** | ***E. coli*** | 6–15 SNPs | 129 | 12 | 9.30 | 5.40 | 15.56 |
| **Hannover** | ***E. coli*** | 16–30 SNPs | 166 | 10 | 6.02 | 3.30 | 10.73 |
| **Hannover** | ***K. pneumoniae*** | 0–5 SNPs | 138 | 85 | 61.59 | 53.27 | 69.29 |
| **Hannover** | ***K. pneumoniae*** | 6–15 SNPs | 68 | 14 | 20.59 | 12.68 | 31.64 |
| **Hannover** | ***K. pneumoniae*** | 16–30 SNPs | 37 | 9 | 24.32 | 13.36 | 40.12 |
| **Hannover** | ***P. aeruginosa*** | 0–5 SNPs | 66 | 38 | 57.58 | 45.56 | 68.76 |
| **Hannover** | ***P. aeruginosa*** | 6–15 SNPs | 54 | 10 | 18.52 | 10.38 | 30.84 |
| **Hannover** | ***P. aeruginosa*** | 16–30 SNPs | 77 | 8 | 10.39 | 5.36 | 19.18 |

The analysis unit was the patient–species–cluster assignment. Nearest genomic distance was defined as the minimum observed SNP distance to an isolate from another patient in the same cluster. When several patients were tied at the minimum distance, the assignment was considered linked if at least one tied nearest co-member had a documented direct or indirect ward contact. The table includes assignments with a nearest co-member 30 SNPs or fewer away; 26 assignments with a nearest within-hospital co-member more than 30 SNPs away are not shown. Proportions are presented with Wilson 95% CIs. CI=confidence interval. SNP=single-nucleotide polymorphism.

**Table S5D. Ward-contact permutation analysis**

| | Copenhagen | | | Hannover | | |
|---|---|---|---|---|---|---|
| | *E. coli* | *K. pneumoniae* | *P. aeruginosa* | *E. coli* | *K. pneumoniae* | *P. aeruginosa* |
| Patient–species–cluster assignments (n) | 732 | 225 | 238 | 515 | 243 | 209 |
| Assignments with proxy ward contact, observed (%) | 4.64 | 6.22 | 5.88 | 14.17 | 20.16 | 17.22 |
| Assignments with proxy ward contact, null median (%) | 0.27 | 0.00 | 0.00 | 0.78 | 4.12 | 1.67 |
| Null distribution, 2.5th percentile (%) | 0.00 | 0.00 | 0.00 | 0.00 | 1.65 | 0.00 |
| Null distribution, 97.5th percentile (%) | 0.82 | 0.89 | 2.10 | 2.33 | 7.41 | 3.83 |
| Permutation *p* | 0.001 | 0.001 | 0.001 | 0.001 | 0.001 | 0.001 |

Proxy ward contact required isolates from two patients to have been sampled on the same ward less than 14 days apart. Cluster labels were randomly permuted across patient–species–cluster assignment rows within each hospital and species, preserving the number of rows assigned to each cluster. Patients with several assignments contributed several rows; unique patient membership within a permuted cluster was not enforced. Results are based on 1000 permutations with a fixed random seed. The empirical one-sided p value used a plus-one correction; the smallest attainable value was 1/1001, reported as 0.001.

**Table S6. Standardisation and model robustness for the hospital comparison**
**Table S6A. Standardisation analyses**

| | Full dataset | Patient-number-matched (1000 draws) | Same Hannover calendar window |
|---|---|---|---|
| Copenhagen patients (n) | 8262 | 5450 | 3229 |
| Hannover patients (n) | 5450 | 5450 | 5450 |
| Crude OR [95% CI] | 1.21 [1.10–1.33] | 1.53 [1.37–1.70] | 1.88 [1.64–2.16] |
| Adjusted OR [95% CI] | 0.96 [0.86–1.07] | 1.23 [1.08–1.39] | 1.49 [1.28–1.73] |
| Crude OR across draws, 2.5th–97.5th percentile | NA | 1.42–1.66 | NA |
| Adjusted OR across draws, 2.5th–97.5th percentile | NA | 1.14–1.33 | NA |

ORs compare the odds of belonging to a local genomic cluster in Hannover with those in Copenhagen. Adjusted models included the presence of any ICU-associated isolate, MDR status, and indicators for the final analytical specimen categories. For patient-number matching, 5450 Copenhagen patients were sampled without replacement in each of 1000 draws and compared with all 5450 Hannover patients. The reported ORs and model-based 95% CI limits are the medians across draws; the additional percentile ranges describe the empirical distribution of the OR point estimates across draws and are not model-based CIs. In the calendar-window analysis, both datasets were restricted to March 2, 2021, to Aug 24, 2022, and clusters and patient-level covariates were reconstructed from isolates within that period. NA=not applicable. CI=confidence interval. ICU=intensive care unit. MDR=multidrug-resistant. OR=odds ratio.

**Table S6B. Model-specification sensitivity**

| | | Patients (n) | OR [95% CI] | *p* |
|---|---|---|---|---|
| **Full dataset** | **M0 Crude** | 13,712 | 1.21 [1.10–1.33] | <0.0001 |
| **Full dataset** | **M1 Hospital + ICU + MDR** | 13,712 | 1.04 [0.94–1.15] | 0.46 |
| **Full dataset** | **M2 Hospital + ICU + specimen composition (no MDR)** | 13,712 | 0.98 [0.88–1.09] | 0.74 |
| **Full dataset** | **M3 Full model** | 13,712 | 0.96 [0.86–1.07] | 0.49 |
| **Full dataset** | **M4 Full model without screening indicator** | 13,712 | 1.09 [0.99–1.21] | 0.08 |
| **Same Hannover calendar window** | **M0 Crude** | 8679 | 1.88 [1.64–2.16] | <0.0001 |
| **Same Hannover calendar window** | **M1 Hospital + ICU + MDR** | 8679 | 1.63 [1.42–1.88] | <0.0001 |
| **Same Hannover calendar window** | **M2 Hospital + ICU + specimen composition (no MDR)** | 8679 | 1.50 [1.30–1.74] | <0.0001 |
| **Same Hannover calendar window** | **M3 Full model** | 8679 | 1.49 [1.28–1.73] | <0.0001 |
| **Same Hannover calendar window** | **M4 Full model without screening indicator** | 8679 | 1.68 [1.45–1.94] | <0.0001 |

Patient-level binomial regression models used membership in any valid local genomic cluster as the outcome. ORs represent the hospital coefficient for Hannover compared with Copenhagen. ICU, MDR, and specimen-category indicators were positive when the corresponding characteristic was present in any isolate from that patient. Screening status overrode the specimen type at the isolate level, although a patient could contribute different isolates from more than one analytical category. M0 included hospital only; M1 additionally included ICU and MDR status; M2 included ICU and all specimen-category indicators but omitted MDR status; M3 was the full model including ICU, MDR status, and all specimen-category indicators; and M4 omitted only the screening indicator from the full model and did not exclude screening isolates. For the calendar-window analysis, clusters and covariates were reconstructed using isolates collected from March 2, 2021, to Aug 24, 2022. *p* values are from two-sided Wald tests. CI=confidence interval. ICU=intensive care unit. MDR=multidrug-resistant. OR=odds ratio.

**Table S7. Sensitivity to a 70-SNP threshold for *P. aeruginosa***

| | Copenhagen | Hannover |
|---|---|---|
| Patients total (n) | 1577 | 1245 |
| Clustered patients at 30-SNP cutoff (n) | 238 | 204 |
| Clustered patients at 30-SNP cutoff (%) | 15.09 | 16.39 |
| Local clusters at 30-SNP cutoff (n) | 64 | 73 |
| Clustered patients at 70-SNP cutoff (n) | 419 | 392 |
| Clustered patients at 70-SNP cutoff (%) | 26.57 | 31.49 |
| Local clusters at 70-SNP cutoff (n) | 86 | 104 |
| Difference in clustered patients (n) | 181 | 188 |
| Difference in clustered patients (percentage points) | 11.48 | 15.10 |
| Difference in local clusters (n) | 22 | 31 |

Global single-linkage components were reconstructed after increasing the *P. aeruginosa* threshold from 30 SNPs to 70 SNPs; the thresholds for *E. coli* and *K. pneumoniae* remained at 30 SNPs in both analyses. A local cluster was defined as the hospital-specific subset of a global component containing isolates from at least two patients. Patient denominators comprise unique patients with *P. aeruginosa* isolates at the respective hospital. Differences were calculated as the result at 70 SNPs minus the result at 30 SNPs; differences between proportions are expressed in percentage points. SNP=single-nucleotide polymorphism.
